# Enabling whole-genome DNA methylation-based classification of central nervous system tumors

**DOI:** 10.1101/2025.09.18.25335800

**Authors:** Jonghoon Lee, Young Seok Ju, Baek-Lok Oh, Seung Ah Choi, Se Hoon Kim, Ji Won Lee, Ji Hoon Phi, Youngmok Jung

## Abstract

**Objectives:** DNA methylation profiling using array-based platforms has proven invaluable for classifying central nervous system (CNS) tumors, especially those with challenging or atypical morphologies. However, existing classification frameworks remain restricted to array-based inputs, interrogating only a subset of CpG sites and limiting diagnostic and prognostic resolution. Whole-genome methylation sequencing methods such as whole-genome bisulfite sequencing (WGBS) and enzymatic methyl-seq (EM-seq) offer near-complete methylome coverage, but their integration into established classifiers is lacking. This study aimed to develop MethylInsight, a web-based platform designed to adapt a widely recognized CNS tumor classification framework to whole-genome data.

**Methods:** MethylInsight converts WGBS and EM-seq signals into array-compatible beta values, enabling compatibility with established classifiers. A Random Forest-based model with logistic regression calibration was trained on 3,905 CNS tumor and control samples spanning 82 tumor subtypes and nine control tissue classes. Performance was evaluated using five-fold cross-validation and internal validation on 22 matched patient samples. The platform also incorporates t-distributed stochastic neighbor embedding (t-SNE) visualizations for contextualizing newly profiled samples against a reference cohort.

**Results:** MethylInsight demonstrated robust classification performance across tumor classes, achieving an area under the ROC curve (AUC) of 0.961, comparable to the DKFZ (0.966) and NM (0.964) classifiers. Cross-validation showed uniformly high accuracy, including for glioblastoma (GBM), a challenging subtype, with sensitivity and specificity of 0.924 and 0.940, respectively. Calibration reduced the estimated error rate from 5.88% to 2.95%. Validation across platforms showed strong concordance, with 21 of 22 paired datasets achieving >80% Pearson correlation, and top-ranked predictions matched for all but three pairs, which still shared overlapping top-two predictions.

**Conclusions:** MethylInsight enables whole-genome methylation data integration into CNS tumor classification, overcoming limitations of array-based methods. By supporting EM-seq inputs and providing calibrated probabilities and intuitive t-SNE visualizations, MethylInsight enhances diagnostic precision and tumor stratification. The platform is freely accessible at https://inocras.methylclassifier.com.

## Introduction

Array-based DNA methylation profiling has demonstrated significant utility in classifying central nervous system (CNS) tumors with challenging or atypical morphology ^1-3^. The Heidelberg Brain Tumor Methylation Classifier^4^ developed at the German Cancer Research Center (DKFZ), has become a widely recognized reference standard in neuro-oncology. This approach examines the targeted CpG sites in the array and enables robust stratification of tumors into distinct subtypes that strongly correlate with genetic alterations, tissue of origin, and clinical outcomes, thereby enhancing diagnostic precision and informing targeted therapeutic strategies ^2,4-6^.

Despite their utility, array-based methods interrogate only a fraction of the methylome, providing insights limited to the targeted CpG sites ^7-9^. In contrast, whole-genome methylation sequencing methods—such as whole-genome bisulfite sequencing (WGBS) and enzymatic methyl-seq (EM-seq)—provide near-complete, unbiased coverage of the methylome. As sequencing becomes more cost-effective, these comprehensive techniques are increasingly accessible, enabling novel biomarker discovery, improved tumor subclassification, and multi-omics integration. In this evolving landscape, adapting established classification frameworks to support whole-genome methylation data enables researchers to enhance tumor stratification with epigenetic insights beyond the limitations of array-based methods.

To this end, we developed MethylInsight, an open-access, web-based platform that extends a well-established central nervous system (CNS) tumor classification framework with whole-genome methylation data. MethylInsight leverages common CpG sites from both array-based and whole-genome (WGBS/EM-seq) datasets by converting these signals into array-compatible beta values. A machine learning (ML)-based predictive model, trained on 3,905 CNS samples spanning across 82 tumor subtypes and nine classes of control tissue, generates calibrated probability estimates, while t-SNE visualizations contextualize new samples within the reference cohort. Five-fold cross-validation and in-house validation on 22 matched patient samples confirm that MethylInsight achieves robust classification performance, expanding the diagnostic utility beyond array-based profiling.

## Methods

### Public Data Sets

Published data were downloaded from GEO under the accession number GSE109381 (HM450K), GSE140686 (HM450K, EPIC), GSE215240 (HM450K, EPIC), GSE130051 (HM450K, EPIC), and GSE54880 (HM450K) ^4,5,10-12^. These datasets collectively encompass over 3,900 CNS tumor and control samples, spanning 82 distinct tumor subtypes and 9 classes of normal tissue across both adult and pediatric cohorts.

### Validation Sample Collection

A total of 22 validation samples were collected from Ajou University Hospital (Suwon, Korea). This study was approved by the Institutional Review Board of Ajou University Hospital (IRB No. 2212-155-1391), and all samples were de-identified prior to analysis. Tumor samples underwent formalin fixation and paraffin embedding (FFPE) by incubation in 10% neutral buffered formalin, followed by standard paraffin embedding procedures. Genomic DNA was extracted from formalin-fixed, paraffin-embedded (FFPE) tumor tissue using the AllPrep DNA/RNA FFPE Kit (QIAGEN, Cat. No. 80234) following the manufacturer’s protocol. Briefly, 10–20 µm sections were deparaffinized and rehydrated prior to lysis in Buffer PKD supplemented with proteinase K at 56 °C for 15 min. Following lysis, the lysate was centrifuged to pellet insoluble material, and the supernatant was processed for RNA isolation, while the retained pellet was further digested and subjected to high-temperature incubation (90 °C for 2 h) to partially reverse formaldehyde crosslinking prior to DNA purification using QIAamp MinElute spin columns. Genomic DNA was eluted in Buffer ATE, and its integrity was verified by agarose gel electrophoresis. Quantification was performed with the Qubit DNA BR assay, and all steps were executed in accordance with the manufacturer’s recommendations.

Following DNA extraction, two platforms were employed for genome-wide DNA methylation profiling: Illumina Infinium MethylationEPIC v2.0 (EPIC-V2) and Enzymatic Methyl-seq (EM-seq). For EPIC-V2, the EPIC-V2 BeadChip assay (Illumina) was performed at Macrogen Inc. (Seoul, South Korea) to generate DNA methylation profiles. In brief, more than 500 ng of FFPE-derived DNA was subjected to bisulfite conversion using the EZ-96 DNA Methylation Kit (Zymo Research Corp.) according to the manufacturer’s recommendations for Infinium assays. The bisulfite-converted DNA was then denatured, neutralized, and hybridized to the EPIC-V2 BeadChip at 48 °C for 16–20 hours. Post-hybridization, the BeadChips were washed, stained, and scanned on an iScan scanner (Illumina), and fluorescence signal intensities were recorded for genome-wide methylation analysis.

For EM-seq, libraries were prepared using a NEBNext Enzymatic Methyl-seq Kit (NEB, #E7120). The input for each library consisted of 100 ng of genomic DNA that had been combined with 1 pg of control pUC19 DNA and 20 pg of control lambda DNA and sonicated to fragments averaging ∼450 bp in length using a Covaris. The protocol for large insert libraries was followed with formamide as denaturing agent, and the libraries were amplified with 5 PCR cycles and sequenced on a NovaSeq 6000 (or NovaSeq X plus) sequencer (Illumina) in paired end 150 bp mode with average depth of coverage of 30x.

### Data Processing

Raw signal intensities for the EPIC-V2 platform were obtained from IDAT files and processed with the methylprep Python package (v1.7.1). To harmonize probe identifiers with those of the HM450K platform, suffixes were removed from probe IDs and duplicated probes were averaged. For example, probes cg00016699_BC21 and cg00016699_BC22 were merged into a single probe, cg00016699, with its beta value defined as the arithmetic mean of the original values.

Paired-end whole-genome methylation sequencing reads from EM-seq were aligned and processed using Bismark (v0.24.0). This workflow enabled the quantification of methylated and unmethylated cytosines across CpG, CHG, and CHH contexts. To convert EM-seq data into beta values that are compatible with array data, the HM450K BeadChip manifest file was employed to map CpG sites to probe IDs. The beta value for each site was calculated as:

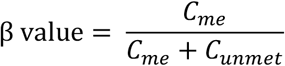

where C_me_ and C_unme_ represent the counts of methylated and unmethylated cytosines, respectively. Missing beta values for any CpG sites were imputed with a value of 0.5. Overall, less than 0.4% of CpG sites were missing (mean: 0.185%, maximum: 0.315%) after conversion to the HM450K format.

### CpG Site Selection

Three publicly available datasets (GSE109381, GSE140686, and GSE215240)^4,5,10^ were integrated to identify common CpG sites across platforms. Autosomal methylation sites were retained by filtering out probes mapping to non-autosomal chromosomes, and the intersection of probes in all datasets yielded 345,314 CpG sites for further analysis.

The CpG sites used for classifier development were refined by implementing a random partitioning strategy using scikit-learn (v1.2.0) ^13^. After randomly shuffling the complete list of CpG sites, the list was segmented into fixed-size subsets. Within each subset, stratified downsampling preserved class proportions, and an independent random forest classifier was trained. Permutation-based feature importance was then assessed by measuring the decrease in class-specific accuracy upon random feature selection.

### ML based Classifier Development

A Random Forest classifier was implemented using the scikit-learn Python package (v1.2.0) ^13^. The initial model was trained on a cohort of 2,801 samples, and its performance was assessed on an independent validation set of 1,104 CNS tumors, with the training and validation splits defined according to the label information in the GSE109381 series metadata ^4,14^. To reduce computational complexity without sacrificing performance, the top 100,000 variable probes were first selected from a total of 428,799 probes. A Random Forest classifier was then used to rank these probes by importance—measured by the decrease in class-specific classification accuracy upon permutation—with the 10,000 most informative features retained for model training. The final model was configured with 10,000 trees and an *mtry* parameter of 100, and to maximize predictive power, MethylInsight was ultimately trained on the combined dataset of 3,905 samples.

### Calibration Model Development

To obtain clinically interpretable probabilities instead of raw scores, the calibration model was trained using five-fold nested cross-validation on the training set. In each fold, one-fifth of the data was held out as an independent validation subset while the remaining four-fifths were used for model training, along with hyperparameter tuning and subsequent performance estimation. As a result, each training sample was independently predicted by a model that had not seen that sample during training. The raw scores obtained from these independent validation folds were then aggregated and used to train an L2-penalized multinomial logistic regression model for score recalibration. The calibration model was implemented using the scikit-learn Python package (v1.2.0) ^13^.

### Unsupervised Analysis

Data visualization was performed using the openTSNE Python package (v1.0.2) ^15^. When newly acquired samples need to be visualized alongside an existing reference cohort, openTSNE enables incremental embedding. Rather than re-computing the entire embedding, it efficiently integrates new data points, preserving the global structure and allowing for rapid visualization. We used the 20,000 most variable CpG sites from 3905 samples from GSE109381 data ^4^, following parameters for t-SNE: *n_components = 2, perplexity = 30, learning_rate = 200, n_iter = 10000, random_state = 42, init = ‘random’, method = ‘barnes_hut’*

### Methylation Class Families

For comparative analysis of existing models, we consolidated methylation subclasses into eight methylation class families (MCFs), as originally described by Capper et al. ^4^. Because the calibrated score of each subclass reflects its probability, we defined the calibrated score of an MCF as the sum of the calibrated scores for all subclasses within that family. The mapping between each methylation subclass and its corresponding MCF is provided in Supplementary Table 1.

### Web Server Design and Implementation

MethylInsight is deployed on Amazon Web Services (AWS) Elastic Beanstalk. When a user uploads data and requests analysis, the workflow is executed in a predetermined order (Figure 1). The frontend was developed using Next.js (v14.2.14) and React (v18.3.1), leveraging HTML5, CSS3, and JavaScript (ECMAScript 2020) to deliver a responsive interface. The backend is powered by Strapi (v4.5.6), which facilitates efficient API management and content handling. High-computation tasks are executed via AWS Lambda, providing on-demand serverless computing for scalability and cost efficiency. The entire application is containerized using Docker to ensure consistent deployment across environments.

**Figure 1.**
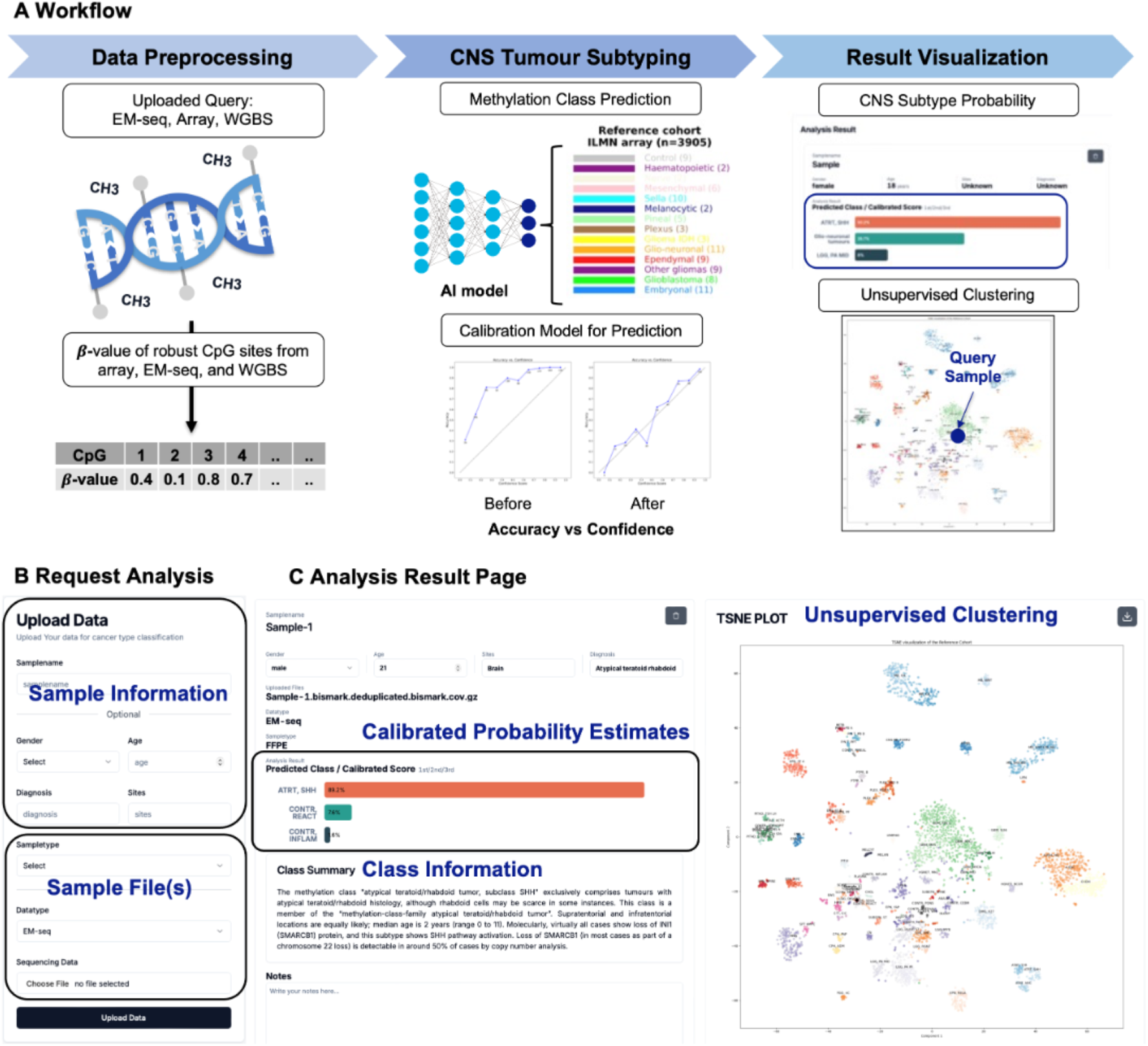
MethylInsight: Workflow, Sample Upload, and Result Visualization. **(A)** Schematic overview of the MethylInsight platform, outlining the preprocessing of whole-genome methylation and array data, followed by classifier prediction and visualization. **(B)** Users can easily initiate analysis by uploading sample information and files. **(C)** On the MethylInsight results page displays, the top three prediction results with calibrated probability estimates, detailed class information, and a t-SNE visualization for comprehensive interpretation.

### Functions

#### Input Requirement

When requesting analysis, the user must complete the upload form with the unified file type(s) and the corresponding data platform (Figure 1B). MethylInsight accepts multiple data types, but input must follow specific formats. For array data, users must provide unprocessed IDAT files containing paired green and red channel data. Filenames should adhere to the format: sampleID_gentrixPosition_Grn (or Red).idat(.gz) and the correct array platform must be selected to call the platform-specific process task. For EM-seq (or WGBS) data, inputs should be final Bismark output files (*.bismark.cov.gz), provided in compressed (.gz) format [9].

## Results

To validate and assess the performance of MethylInsight we compared it with two well-established classifiers (NM and DKFZ). We evaluated the performance of the MethylInsight classifier using the same training dataset of 2,801 samples and an independent validation set of 1,104 samples ^4,16^. The area under the ROC curve (AUC) was computed for all three model(s), yielding comparable values of 0.966 for the DKFZ classifier, 0.964 for the NM classifier, and 0.961 for the MethylInsight classifier (Figure 2B).

**Figure 2.**
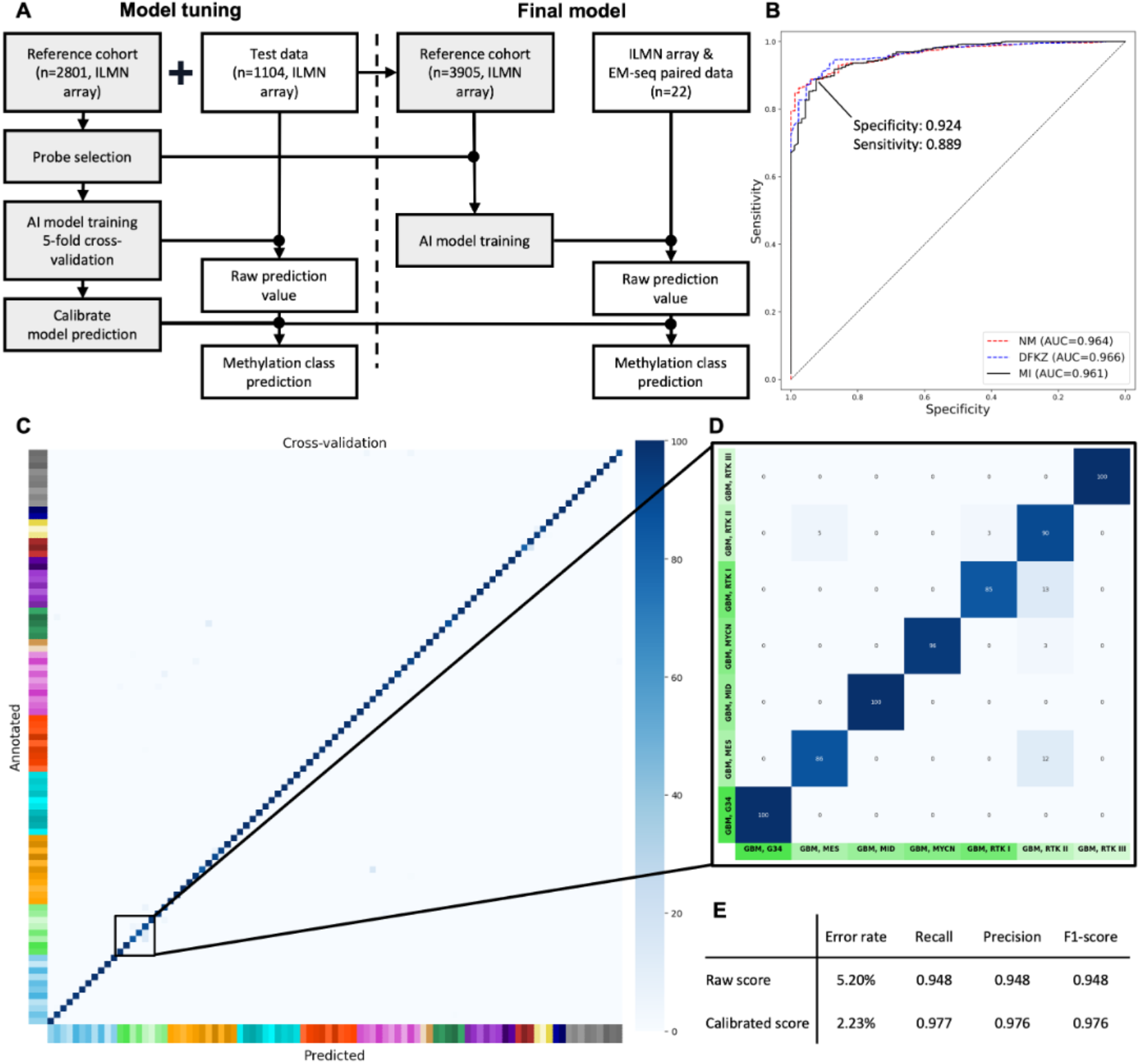
Development and performance of MethylInsight. **(A)** Workflow for classifier development, illustrating principal components (gray) and platform-specific processing steps (white). Features for the random forest were selected based on importance scores. Raw scores from decision trees were calibrated to estimated probabilities. After initial evaluation with a training–test split, the classifier was retrained on the entire GSE109381 dataset to create the final model, which was then validated across different methylation profiling platforms. **(B)** The receiver-operating characteristic (ROC) curves for the NM, DKFZ, and MethylInsight classifiers were generated using 1104 samples. The area under the curve (AUC) values for the DKFZ, NM, and MU classifiers are 0.966, 0.964, and 0.961, respectively. The optimal threshold, as determined by the Youden index, is 0.944, which corresponds to a specificity of 0.924 and a sensitivity of 0.889. When applying a threshold of 0.999, the classifiers achieve a perfect specificity of 1 and a sensitivity of 0.671, while a threshold of 0.9 yields a specificity of 0.87 and a sensitivity of 0.911. **(C)** Confusion matrix from fivefold cross-validation of the final model, showing the classification of 3,905 samples assigned to the class with the highest calibrated probability. **(D)** Enlarged view of the MCP_GBM class. **(E)** Improvement in prediction accuracy of the random forest classifier, constructed from 3,905 biologically independent samples, achieved through score calibration.

Classifier development proceeded in two main steps (Figure 2A). The final model was constructed based on the hyperparameters, selected CpG sites, and a fitted logistic regression calibration model obtained during the model tuning process. The model’s performance was subsequently assessed via cross-validation. The resulting confusion matrix confirmed that the final model achieved uniformly high accuracy across all classes (Figure 2C). Notably, for the GBM class—well recognized for its classification difficulty ^4^—the model achieved a sensitivity of 0.924 and a specificity of 0.940 (Figure 2D). Furthermore, the Random Forest classifier exhibited an estimated error rate of 5.88% when using raw scores, which decreased to 2.95% after calibration (Figure 2E).

### Cross-Platform Performance Validation

Our service specializes in methylation class classification through an integrated model that comprises two main components: data preprocessing and classification. Upon data upload, each sample is processed according to its data type, and the system generates calibrated predictions of the methylation classes. When users upload their data, the system processes the sample according to its data type and then predicts methylation classes with calibrated scores (Figure 3A and 3B). The top three classes are presented independently of the predicted score (Figure 1C).

**Figure 3.**
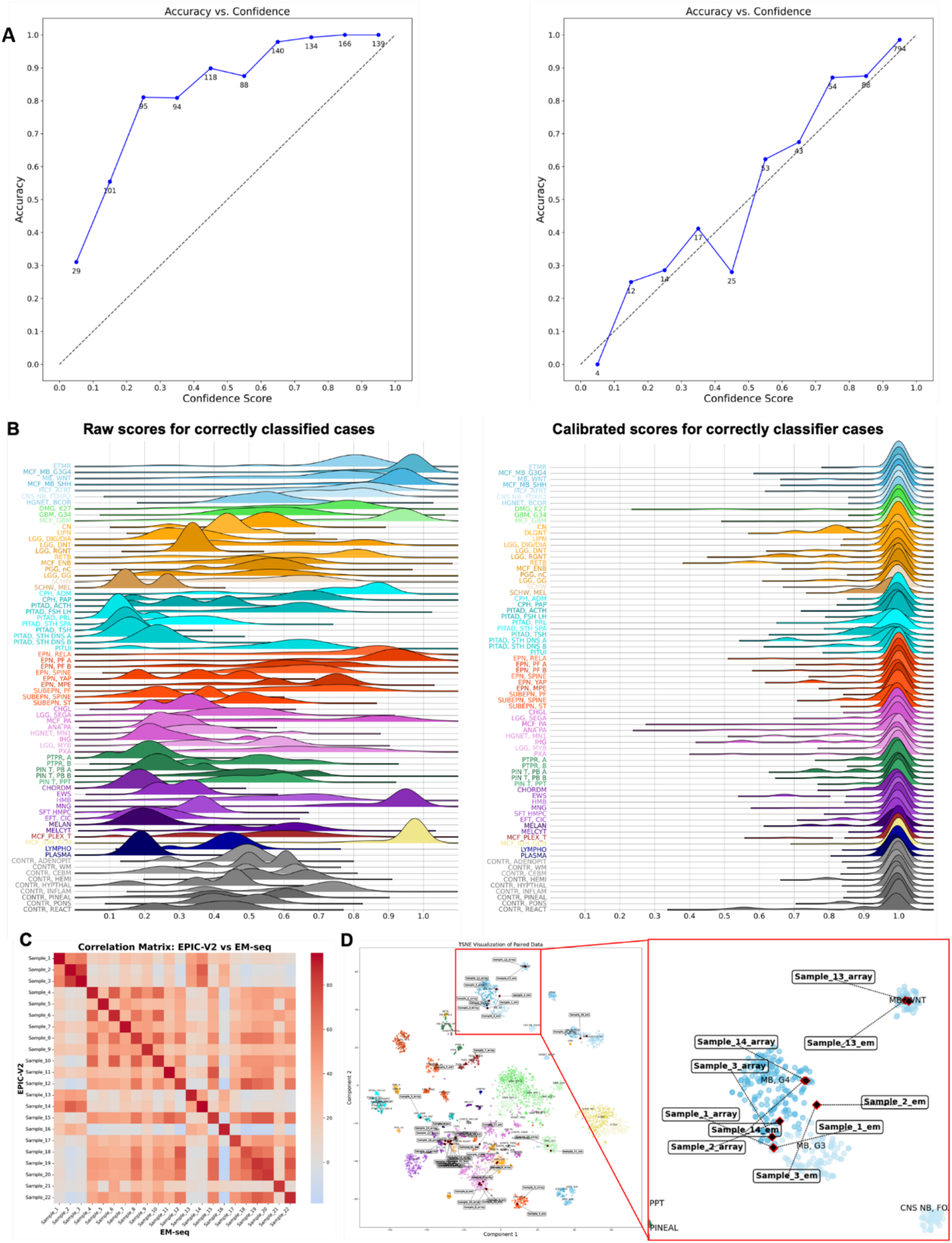
Comparative analysis of raw and calibrated scores and EM-seq and EPIC-V2 platforms. **(A)** Accuracy versus confidence score: after calibration, confidence scores more closely align with true accuracy. **(B)** Density plots of distribution of raw and calibrated scores for correctly classified samples: score calibration reduces inter-class variability. **(C)** t-SNE plot of paired samples analyzed on both EM-seq and EPIC-V2, each pair mapped to the same coordinates, demonstrating cross-platform consistency. **(D)** Heat map showing pairwise Pearson correlation of the 22 paired datasets: processed exhibit high within-sample correlation despite platform differences.

To verify the consistency of prediction results across different platforms, we conducted additional experiments. First, to assess whether the processed data exhibited consistent beta-values at individual CpG sites, Pearson correlation coefficients were computed for each paired dataset following the preprocessing procedures described in the Materials and Methods section. Of the 22 paired datasets, 21 exhibited correlation coefficients exceeding 80% (with the lowest at 76.1%), yielding an average correlation of approximately 88% (Figure 3C). Moreover, t-SNE visualization demonstrated that the paired datasets clustered closely with the reference cohort (Figure 3D).

Furthermore, comparison of the MethylInsight classifier’s prediction outcomes showed that, with the exception of three pairs, the top-ranked predicted class was identical across datasets and the predicted scores were comparable. Even for the three unmatched pairs, the same class appeared within the top two predictions (Table 1). The paired datasets were also analyzed using the DKFZ white (classifier v12.8) ^4^, and in comparison, to the DKFZ results, four inconsistent outcomes were observed (Table 1). For instance, Sample 1 and Sample 2—representing Medulloblastoma groups 3 and 4 (MB, G3 and MB, G4), which are frequently misclassified due to their histological and biological similarity ^4,11^.

**Table 1.** Comparison of Prediction Results Against DKFZ.

| Sample Name | Prediction Result on EM-seq | Prediction Result on EPIC-V2 | Consistency with DKFZ Prediction | DKFZ Result on Illumina Array |
| --- | --- | --- | --- | --- |
| Sample1 | MB, G3 (0.994), MB, G4 (0.005) | MB, G3 (0.996), MB, G4 (0.003) | O | MB, G4 (0.673) |
| Sample2a | MB, G4 (1.0), MB, G3 (0) | MB, G4 (1), MB, G3 (0) | X | MB, G4 (0.961) |
| Sample2b | MB, G4 (0.948), MB, G3 (0.052) | MB, G4 (0.989), MB, G3 (0.006) | X | MB, G4 (0.967) |
| Sample3 | LGC, DNT (0.997), HG (0.001) | LGC, DNT (0.997), LGC, PAP (0.002) | O | LGC, DNT (0.995) |
| Sample4 | CPH, DAM (1), CPH, PAP (0) | CPH, DAM (1), CPH, PAP (0) | O | CPH, ADM (1) |
| Sample5 | LCG, PA PF (0.536), LGC, PA MID (0.439) | LCG, PA PF (0.546), LGC, PA MID (0.45) | O | LGC, PA PF (0.577) |
| Sample6 | PLEX, AD (0.998), PLEX, ADP (0.001) | PLEX, PED (1) | O | PLEX, PED (1) |
| Sample7 | LEC, PA, PF (0.891), CONTR, REACT (0.095) | CONTR, REACT (0.583), LGC, PA MID (0.369) | X | LGC, PA PF (0.569) |
| Sample8 | EPN, RELA (1), EPN, YAP (0) | EPN, RELA (1), EPN, YAP (0) | O | EPN, RELA (1) |
| Sample9 | LCG, PA PF (1), LGC, PA MID (0) | SCHW, MEL (0.359), DLGNT (0.146) | O | EPN, RELA (0.222) |
| Sample10 | SCHW, MEL (0.519), DLGNT (0.006) | CONTR, REACT (0.392), CONTR, REACT (0.387) | Not supported | VM, TMH NEG (0.997) |
| Sample11 | CONTR, REACT (0.938), CONTR, REACT (0.042) | MB, WNT (1), MB, G3 (0) | O | EPN, RELA (0.222) |
| Sample12 | MB, WNT (1), MB, G3 (0) | MB, G4 (1), MB, G3 (0) | O | MB, WNT (0.968) |
| Sample13 | MB, G4 (1), MB, G3 (0) | SCHW, MEL (0.93), DLGNT (0.015) | X | MB, G4 (0.963) |
| Sample14 | SCHW, MEL (0.852), SCHW (0.023) | MB, SHH CHL (A) (1), SHB, MB (0) | O | VM, TMH NEG (0.5830) |
| Sample15 | MB, SHH CHL (A) (1), SHB, MB (0) | INF (1), INF (0) | O | MB, SHH CHL (0.971) |
| Sample16 | INF (1), INF (0) | CONTR, REACT (0.967), CONTR, REACT (0.962) | X | CPT, GERM A (0.8015) |
| Sample17 | CONTR, MELON (0.474), CONTR, REACT (0.426) | SCHW, MEL (1.005) | O | EPN, RELA (0.0494) |
| Sample18 | SCHW, MEL (0.994), ERKT (1.005) | SCHW, MEL (0.994), HMB (0.026) | X | CPT, GERM A (0.41971) |
| Sample19 | SCHW, MEL (0.831), MB (0.097) | SCHW, MEL (0.964), HMB (0.031) | O | VM, TMH NEG (0.17633) |
| Sample20 | SCHW, MEL (0.956), CHORD (0.044) | DMG, K27 (1), LGC, RGNT (0) | X | DMG, K27 (1) |
| Sample21 | DMG, K27 (1), LGC, RGNT (0) | MNG (0.583), SFT HMB (0.0331) | O | MNG (0.28233) |

Note. Consistency with DKFZ prediction is marked as O (consistent) X (inconsistent), or Not supported.
<sup>1</sup>'O' indicates that the classification results across the different classifiers converge on the same MCF class, whereas 'Not supported' denotes methylation classes that are present exclusively in DKFZ v12.8 and are not recognized by the Methyllnsight classifier.
<sup>2</sup>Detailed information for each class abbreviation is provided in Supplementary Table 1.

To investigate the reasons for the inconsistent prediction results between the two models, we performed t-SNE plots on Medulloblastoma-specific data extracted from several datasets (GSE109381, GSE130051, GSE54880) ^4,11,12^ for Sample 1, Sample 2a, and Sample 2b, enabled a combined t-SNE plot (Supplementary Figure 1). The visualization of t-SNE revealed that Sample 1 grouped with the MB, G3 group, while Sample 2a and Sample 2b were placed in the MB, G4 group, in alignment with the results of the MethylInsight classifier rather than those of the DKFZ classifier. Moreover, since Sample 2a and Sample 2b originate from the same sample, they should produce identical classifications (MB, G4); however, the DKFZ classifier provided inconsistent results for these cases. Additionally, two other cases exhibited extremely low confidence scores (0.222 and 0.0404 for Samples 11 and 17, respectively). These findings collectively indicate that MethylInsight produces outcomes in strong agreement with the well-established DKFZ model for EM-seq data, while offering enhanced accuracy in cases that are inherently challenging to distinguish.

### Unsupervised Clustering Result Visualization

To visualize sample data quickly and consistently, we used the openTSNE module along with a consistent CpG sites for class embedding ^15^. Once the sample data were processed, they were utilized for both classification and visualization. Users can verify the classification results through visualization by viewing the images, and by comparing the t-SNE results with the predicted outcomes, they can validate predictions ^4^.

When a user requests an analysis, MethylInsight provides both the analysis results and a corresponding t-SNE image. t-SNE, which handles unbiased results, offers the advantage of enabling users to assess the reliability of the classifier’s outcomes by comparing the classification results with the t-SNE image ^4^. Moreover, the t-SNE image is generated using the openTSNE module, which ensures that the embedding results for the reference cohort remain consistent, even as new samples are added ^15^. It makes producing rapid and consistent t-SNE outcomes.

## Conclusions

Our study presents MethylInsight, an open-access, web-based platform that expands CNS tumor classification beyond conventional array-based methylation profiling by supporting whole-genome methylation sequencing data such as EM-seq (or WGBS). The system demonstrates robust performance, evidenced by comparable AUC values and low error rates relative to established DKFZ and NM classifiers, while additionally providing calibrated probability estimates and intuitive t-SNE visualizations for prediction validation ^4,16^.

A key advantage of MethylInsight is its ability to accept EM-seq data, a format that is not typically supported by conventional classifiers developed for array-based data. Array-based methods, such as the Heidelberg Brain Tumor Methylation Classifier ^4^, focus on a predetermined set of CpG sites and are therefore inherently limited in scope ^7^. In contrast, whole-genome sequencing approaches offer near-complete, unbiased coverage of the methylome, allowing for the discovery of novel biomarkers and more nuanced tumor subclassification ^17,18^. By converting EM-seq signals into array-compatible beta values, MethylInsight bridges this gap and expands the diagnostic capabilities beyond those of traditional array-based methods.

Moreover, the current implementation of MethylInsight converts EM-seq data into an array-compatible format in a relatively straightforward manner. While this approach facilitates integration with existing analysis pipelines, it does not fully capitalize on the rich, global epigenetic information present in EM-seq data. Future iterations of MethylInsight will aim to directly harness the intrinsic properties of whole-genome methylation data. For instance, advanced analytical modules could be introduced to perform unsupervised clustering, integrate multi-omics data, or identify novel epigenetic markers—thereby overcoming the inherent limitations of array-based profiling ^19,20^.

In summary, MethylInsight exemplifies a significant advancement in CNS tumor diagnostics by supporting data formats that conventional models cannot handle. Its capacity to integrate and visualize whole-genome methylation data paves the way for more comprehensive analyses and improved tumor stratification. Moreover, its deployment on AWS Lambda ensures high scalability and rapid runtime, achieving processing costs below $0.1 per sample, making it economically feasible for large-scale clinical use. As sequencing technologies continue to evolve, platforms like MethylInsight will be instrumental in translating complex epigenetic landscapes into clinically actionable insights.

## Data Availability

MethylInsight is freely accessible to the research and clinical communities (https://inocras.methylclassifier.com), supporting transparency and broad adoption.

## Acknowledgments

We thank the contributors of publicly available datasets, including those deposited in GEO, which were instrumental to the development and validation of this study. We also acknowledge Ajou University Hospital for providing matched CNS tumor samples. MethylInsight was developed and hosted by Inocras Inc., with infrastructure and engineering support from its San Diego and Seoul-based teams. We also thank the anonymous reviewers for their thoughtful feedback and suggestions.

## Ethical Considerations

The validation component of this study was approved by the Institutional Review Board of Ajou University Hospital (IRB No. 2212-155-1391). All samples were de-identified prior to analysis.

## Consent to Participate

This study involved archival, de-identified biospecimens and was determined by the IRB to be exempt from informed consent requirements.

## Declaration of Conflicting Interests

Several authors (J.L., Y.S.J., B.-L.O., and Y.J.) are employees of Inocras Inc., the company that developed the MethylInsight platform evaluated in this study. The remaining authors declare no competing interests.

## Funding

This work was supported in part by the National Science Foundation (NSF: #1636933 and #1920920) and by internal research and development funding from Inocras Inc.

## Notes

### Competing Interest Statement

JL, JSJ,BO, YJ are all employees of Inocras

### Author Declarations

A total of 22 validation samples were collected from Ajou University Hospital (Suwon, Korea). This study was approved by the Institutional Review Board of Ajou University Hospital (IRB No. 2212-155-1391), and all samples were de-identified prior to analysis. Public Data Sets Published data were downloaded from GEO under the accession number GSE109381 (HM450K), GSE140686 (HM450K, EPIC), GSE215240 (HM450K, EPIC), GSE130051 (HM450K, EPIC), and GSE54880 (HM450K) 4,5,10-12. These datasets collectively encompass over 3,900 CNS tumor and control samples, spanning 82 distinct tumor subtypes and 9 classes of normal tissue across both adult and pediatric cohorts.

